# Unravelling the population structure and transmission patterns of *Mycobacterium tuberculosis* in Mozambique, a high TB/HIV burden country

**DOI:** 10.1101/2022.02.01.22270230

**Authors:** B. Saavedra, M.G. López, Á. Chiner-Oms, A.M. García, I Cancino, M. Torres-Puente, L. Villamayor, C. Madrazo, E. Mambuque, VG. Sequera, D. Respeito, S. Blanco, O. Augusto, E. López-Varela, AL. García-Basteiro, I. Comas

**Author notes:** Both authors contributed equally as senior authors (AL. García-Basteiro, I. Comas).

## Abstract

Genomic studies of *Mycobacterium tuberculosis complex* (MTBC) might shed light on the dynamics of its transmission, especially in high-burden settings, where recent outbreaks are embedded in the complex natural history of the disease. We applied Whole-genome sequencing (WGS) to characterize the local population of MTBC, unravel potential transmission links and evaluate associations with host and pathogen factors.

**Methods:** A one-year prospective study was conducted in Mozambique, a high HIV/TB burden country. WGS was applied to 295 positive cultures. We combined phylogenetic, geographical and clustering analysis, and investigated associations between risk factors of transmission.

**Findings:** A significant high proportion of strains were in recent transmission (45.5%). We fully characterized MTBC isolates by using phylogenetic approaches and dating evaluation. We found two likely endemic clades, comprised of 67 strains, belonging to L1.2, dating from the late XIX century and associated with recent spread among PLHIV.

**Interpretation:** Our results unveil the population structure of MTBC in our setting. The clustering analysis revealed an unexpected pattern of spread and high rates of progression, suggesting the failure of control measures. The long-term presence of local strains in Mozambique, which were responsible for large transmission among HIV/TB coinfected patients, hint at possible coevolution with sympatric host populations and challenge the role of HIV in TB transmission.

**Funding:** Ministry of Enterprise and Knowledge (Government of Catalonia & European Social Fund, AGAUR fellowship); European Research Council (ERC) European Union’s Horizon 2020.

## Introduction

Tuberculosis (TB) remains the deadliest and one of the most prevalent infectious diseases worldwide.^1^ More than twenty years after the introduction of molecular tools in TB, it is now undoubted that unraveling the transmission dynamics of local epidemics is essential to tackle the ongoing spread.^2^ A complete understanding of who transmits, where, how and why, is essential for designing effective control interventions.^3^

The development of Whole-genome Sequencing (WGS) techniques is transforming the classical vision of the infection and decoding links unreachable by basic epidemiology or traditional genotyping.^4^ However, WGS is still scarcely applied in high-burden settings, where recent outbreaks are embedded in the complex natural evolution of the disease.^5^

On one hand, high-resolution genomic data allows for accurate cluster analysis to investigate outbreaks and decrypt transmission profiles. By using the SNPs cut-off approach, we can delineate transmission clusters with a resolution not seen before.^4^ On the other hand, the study of the origin and genetic diversity of local strains also helps to survey the evolutionary history of the *Mycobacterium tuberculosis complex* (MTBC) population and the potential variations between lineages.^5,6^

Phylogeographical methods have identified different lineages and sub-lineages of MTBC with differential distribution worldwide.^7^ This striking difference in geographic distribution has led to hypotheses as to why some are more widespread than others. Whereas Lineage (L) 4 is the most ubiquitous globally, L1 and L3 have been described as endemic to several high-burden regions in Asia and Eastern Africa.^8^ Although part of those differences can be explained by historical contingency^9^, host-pathogen coevolution is assumed to play a decisive role.^10^ In San Francisco, In San Francisco, there seemed to be a preference of transmission between lineages and hosts with the same geographic origin.^11^ Furthermore, some works suggest that this link between human populations and lineages from the same geographic origin breaks down with HIV status, suggesting a role for coevolution at immune system level.^11^ However, identifying the causes why a genotype succeeds locally remains a challenge and there is limited information on the interaction with other factors, such as HIV infection.

All the insights we are gaining from WGS data should be transferred to high-burden settings and used to define effective public health interventions. Mozambique is a country with one of the highest rates of TB and HIV/TB coinfection^1^, but information on MTBC population structure is scarce in the region.^12,13^ This is, to our knowledge, the first comprehensive population-based study in Mozambique on the application of WGS to characterize local strains, unravel likely transmission links, and evaluate possible associations with host and pathogen factors.

## Methods

### Study design and study population

This was a one-year prospective surveillance-based study (TOSSE study) implemented from August 2013 to 2014. It was conducted in the District of Manhiça, Maputo province, a semi-rural area in Southern Mozambique, 80 kilometers north of the capital, with an estimated population of 186,241 inhabitants at the time of the study (2013-2014)^14^, living in an area of 2,373 km.^15^ This is high TB/HIV burden area with a long history of high TB transmission.^16,17^

Presumptive TB adults, without history of previous TB treatment, presenting with TB compatible symptoms^18^ (no time criteria if HIV-positive), who attended any of the health units belonging to Manhiça District Hospital’s catchment area, were consecutively enrolled.^19^ Only participants with confirmed MTBC in culture were included in the analysis.

We consecutively enrolled participants who met the following criteria: presumptive TB-infected adults, without history of previous TB treatment, presenting TB-compatible symptoms (no time criteria in the case of people living with HIV), attending any of the health units belonging to Manhiça District Hospital’s catchment area.

### Diagnostic procedures

Participants provided two sputum samples at time of diagnosis. Extra-pulmonary specimens were collected at hospital level. Diagnostic tests were performed at the *Centro de Investigação em Saúde de Manhiça* (CISM) - Biosafety level 3 (BSL3) laboratory, which is subject to external quality control and is ISO certified.

#### Ziehl-Neelsen (ZN)

Smear microscopy was done by Ziehl-Neelsen staining. Results were reported as negative or on a scale of positive grades according to international standards.^20^

#### Xpert MTB/RIF (Xpert)

Raw samples were tested according to the manufacturer instructions.^21^ Invalid results were excluded. The semiquantitative results for Xpert fell under the following categories: very low, low, medium, or high.

#### Solid and liquid culture

Samples with positive Xpert results were cultured. Remnant raw samples were decontaminated by Kubica method^23^ and resuspended. Afterwards, 500 microliters were inoculated into Mycobacteria Growth Indicator Tubes (MGIT) liquid medium and incubated in the Bactec MGIT 960 mycobacterial detection instrument (Becton Dickinson Microbiology System, BD, USA). Additionally, 200 microlitres were cultured in BD Lowenstein Jensen solid medium. After 42 days (for liquid culture) or 8 weeks (for solid culture) without growth, samples were classified as negative. In case of positive results, MTBC was confirmed using ZN staining and BD TB Identification test (Becton Dickinson Microbiology System, USA). Isolates were stored at -80ºC.

### Sequencing library construction and bioinformatics pipeline

Culture isolates were shipped to the BSL3 laboratory of *the Instituto de Biomedicina de Valencia* (Spain). After inactivation, samples were used to prepare WGS libraries. Genomic libraries were constructed with the Nextera XT Sample Preparation kit (Illumina Inc., San Diego, CA) according to the manufacturer’s protocol^24^, with 12 cycles for indexing PCR. WGS was carried out in the MiSeq platform (2×300 cycles paired-end run; Illumina).

Sequence analysis was performed following a validated, previously-described bioinformatics pipeline.^25^ Briefly, FASTQ files were preprocessed with *fastp*^26^ in order to trim poor quality bases and potential sequencing errors. Later, in order to reduce likely contaminant reads, we classified and filtered those that did not belong to the MTBC using Kraken.^27^ Samples with less than 90% of MTBC reads were discarded for posterior analyses. After this filtering, reads were mapped against the MTBC most probable ancestral genome^28^ using the BWA-mem algorithm.^29^ Later, we discarded reads with ambiguous mapping based on the BWA MAPQ score (keep those with MAPQ=60) as well as potential duplicate reads by using *picard* tools. Samples with genomic coverage <90% were discarded. The variant calling was performed by a combination of SAMtools and VarScan.^30^

In order to avoid mapping errors and false SNPs, we kept variants that (i) were supported by at least 20 reads, (ii) were found in a frequency of at least 0.9, (iii) were not found inside detected indel areas, or (iv) were found in areas of high accumulation of variants (more than three variants in a 10-bp defined window). Variants were annotated using SnpEff. ^31^ Variants present in PE/PPE genes, phages, or repeated sequences were not considered. With these high-quality variants detected, we generated the alignment. Samples with at least two phylogenetic variants at frequency >10% were classified as mixed infection cases.

### Phylogenetics and geographical analysis

Maximum-likelihood phylogeny was constructed with IQ-TREE^32^ under the General Time Reversible (GTR) model of evolution, with a bootstrap of 1000 replicates and using the *-fconst* option for accounting for invariant sites. Known drug-resistant positions were not considered for generating the phylogeny as they are highly homoplastic. Later, we aimed to define the population structure of closely related strains by using the fast hierarchical Bayesian Analysis of Population Structure (BAPS) algorithm implemented in R library *fastbaps*. BAPS groups were defined under the second level of clustering hierarchy.^33^

#### Geographic origin of the clades from Mozambique (MZ)

In order to unravel the potential geographic origin of the BAPS groups, we used the RASP program.^34^ This is based on both, Bayesian and parsimony approaches, and aims to estimate the ancestral geographic origin. It requires a phylogeny and the geographic origin of the tips as input. We reconstructed a phylogeny combining MZ isolates and 8,263 genomes representative of the MTBC global genetic diversity. We marked MZ strains as having their geographical origin in Mozambique, and strains from other datasets as having non-Mozambique origin. We coupled the Statistical-Dispersal Vicariance Analysis (S-DIVA) with a Bayesian Binary MCMC. For each analysis, we run 5 MCMC chains with 500000 cycles.

RASP output estimates the likely origin of ancestral nodes. We manually defined likely endemic clades as follow: i) they consisted in 1 or more BAPS, ii) the clade contained more than 10 samples from Mozambique and constituted >60% of the total clade, iii) the origin of the most recent common ancestor (MRCA) obtained with RASP was Mozambique (>80% of probability). Non-endemic clades were defined as those in which i) the clade contained more than 10 samples from Mozambique but constituted <60% of the total clade, and ii) the MRCA was not Mozambican. Unknown clades were those which did not meet any of the previous criteria.

### Estimation of times

Dating analysis was performed by applying the Bayesian inference implemented in BEAST2 v2.6.5^35^ A multifasta file was generated for the three major lineages (L1, L2 and L4). We used them as partitions in *beauti*. A GTR substitution model was defined with gamma count = 4 and empirical nucleotide frequencies. A strict clock model was selected; since we do not have tip or node calibration, we fixed different clock rates for each lineage (L1: 1.57 × 10^−7^; for L2: 4.1 × 10^−8^; L4: 3.79 × 10^−8^), according to *Menardo et al, 2019*.^36^ We evaluated exponential and constant coalescent population models, and selected the former, since we found evidence against the constant model (95% HPD of the rate growth not including 0). We chose an exponential prior for the population size (mean = 1, offset = 0), and for the exponential growth rate prior, we used the standard Laplace distribution (mu = 0.001, scale = 30.7). We corrected the xml files to specify the number of invariant sites as indicated here: https://groups.google.com/g/beast-users/c/QfBHMOqImFE. We ran two runs with 2 × 10^7^ steps-long chains, sampling every 2000 steps and removing the initial 10% as burn-in. We evaluated that the mixing and the estimated sample size (ESS) of the posterior and of all parameters were larger than 200, with Tracer 1.7.1.^37^ We used *logcombiner* to combine the tree files from the independent runs and ggtree^38^ R package to annotate and plot the trees.

Lastly, by combining results from molecular dating and the likely ancestral geographical origin from RASP, we aimed to assess for how long previously defined clades have been circulating within the country. We calculated the median and 95%HPD (Highest Posterior Density interval) for MRCAs with origin in Mozambique for all clades, and we compared the average of the median years of introduction in the country among endemic and foreign clades.

### Identification of recent transmission events

Transmission was evaluated by clustering analysis from pairwise distance. We employed the strict relatedness cut-off of 5 SNPs for describing recent transmission^39^ and we evaluated up to 10 thresholds for broad cluster definition. Clusters obtained from Mozambique (estimated incidence rate (IR): 551/100,000 in 2014) were compared to published transmission data (from population-based studies) from settings with different TB burden (Malawi, IR 2009 243/100,000; Valencia, Spain, IR 2015: 9.13/100,000). From available datasets, we chose annual data from 2009^40^ and 2015 (manuscript under revision) respectively, based on the largest datasets close to 2013/4 and to have comparable timespan.

### Statistical analysis

Statistical analyses were performed using the R statistical language (R version 3.5.2, The R Foundation for Statistical Computing Platform). For the phylogenetic analysis, branch lengths were extracted using the *geiger*^41^package. The *fastbaps*^42^ package was applied for the BAPS algorithm. For transmission analysis, transmission events were identified by using *ape* and *adegenet* packages. *Tidyverse, tigerstats, naniar, epiR, purr, broom, ggplot and parameters*,were used to visualize, describe and analyze epidemiological data.

We aimed to explore the association of being in transmission (dependent variable) with a range of collected risk factors (Table 1). The non-parametric Fisher’s exact test was used to identify differences in the distribution of independent variables. We hypothesized that transmission patterns would differ depending on the origin of clades (endemicity). Saturated logistic regression models were used to estimate mutually adjusted odds ratio (OR). Covariates with p-value<0.2, passed for the adjusted analysis. Age, sex and HIV status were chosen as *a priori* related risk factors irrespective of univariate associations. A backward strategy was used to finalize the model. Afterwards, considering the large prevalence of HIV-positive patients in our dataset, and that this unbalanced distribution might confound our findings, we stratified the analysis by HIV status (Table 2).

**Table 1.**
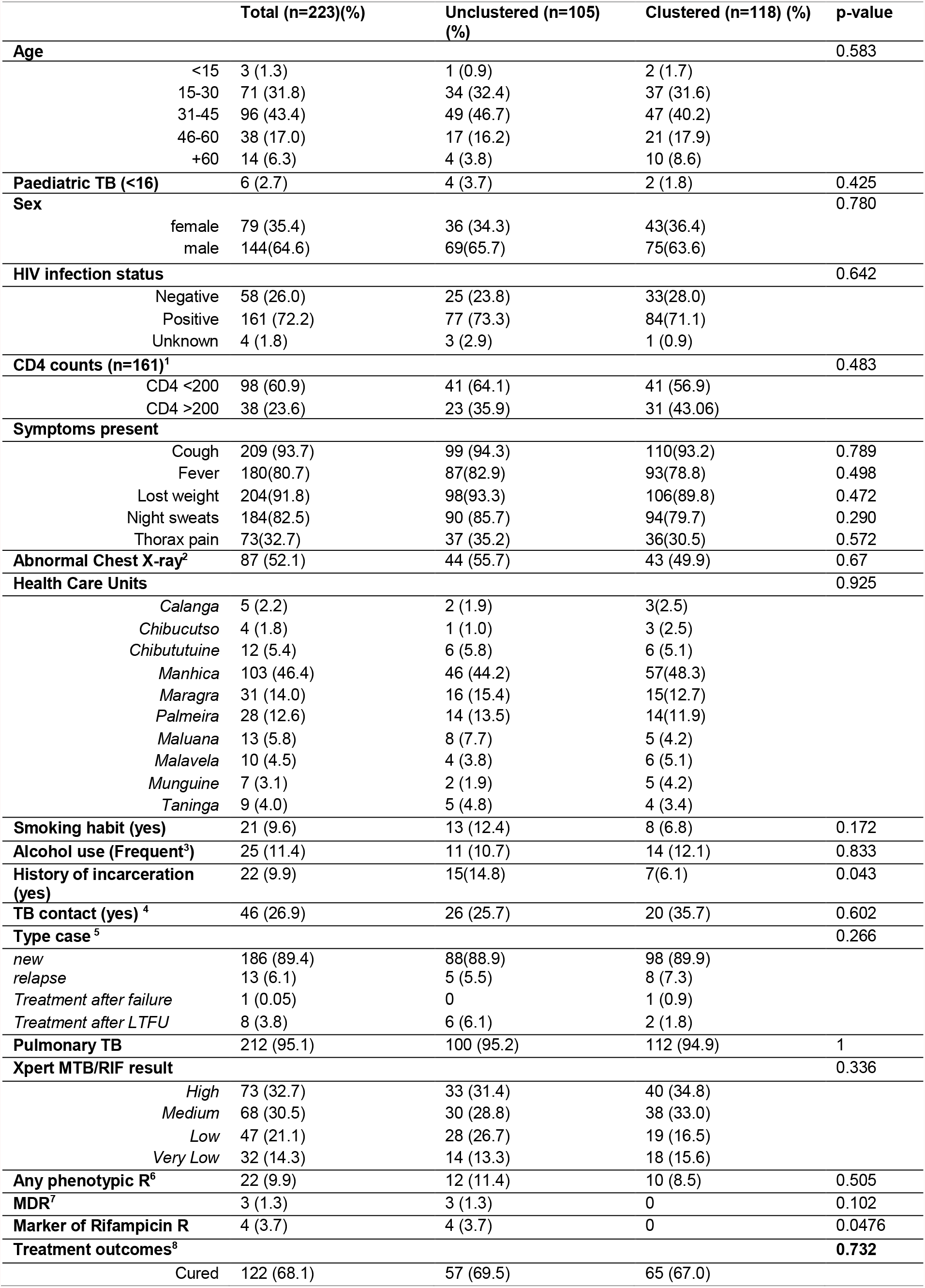
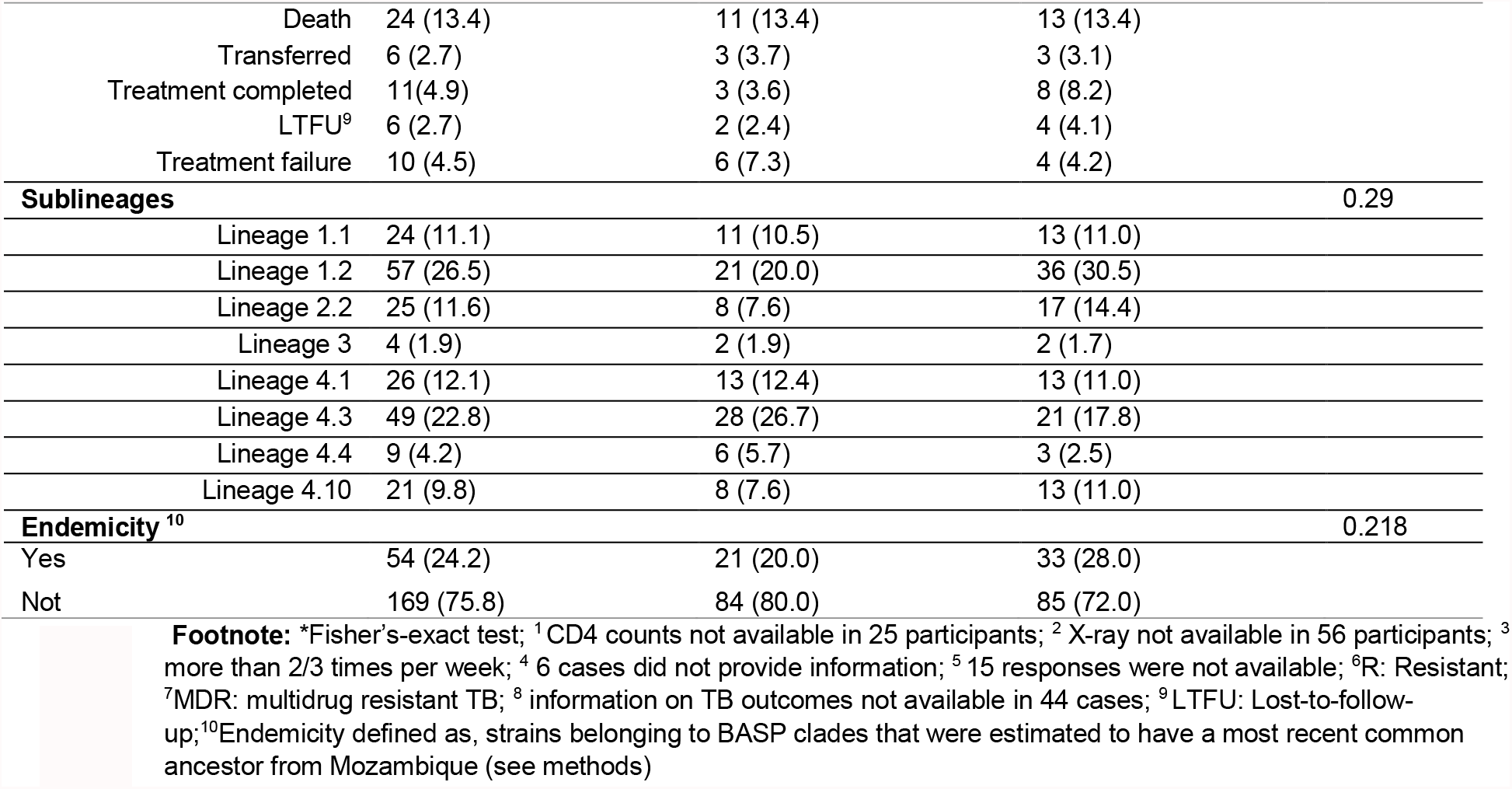
Descriptive analysis of exploratory covariates stratified by clusters of transmission.

**Table 2.**
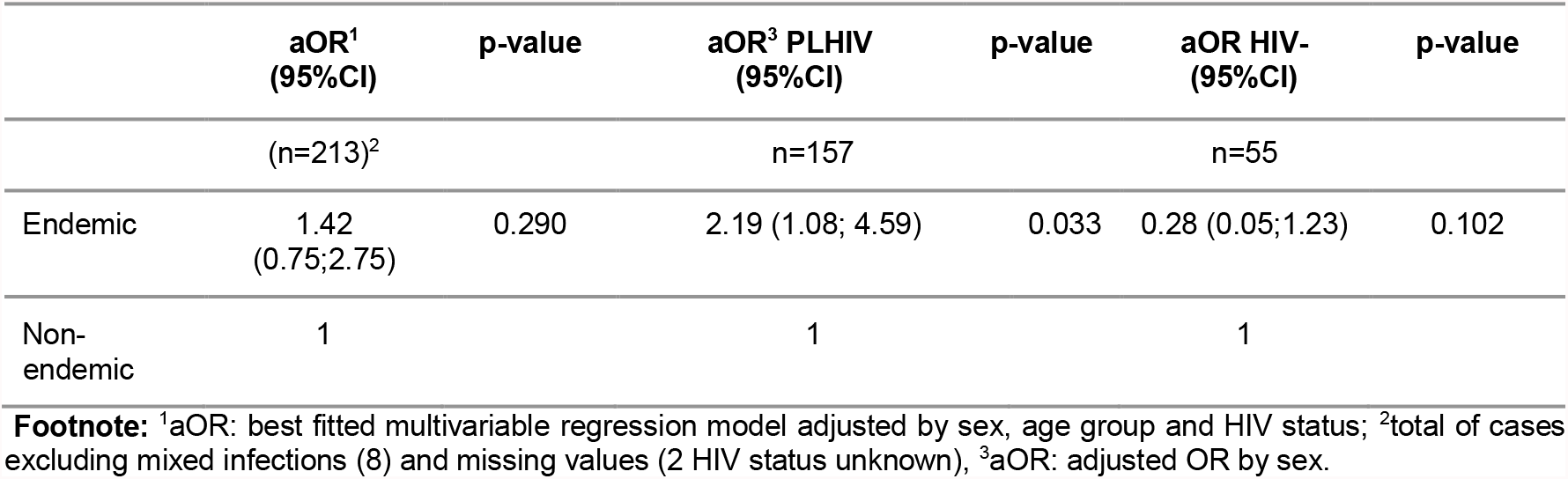
Multivariable logistic regression results on the association of endemicity with clustering as independent variable.

### Ethics

The CISM’s Internal Scientific and Internal Bioethics Committees and the National Bioethics Committee (Maputo, Mozambique) waived ethical approval for this work (Reference: 199/CNBS/13). Written informed consent was provided by all study participants

## Results

### 1. Overall population structure of MTBC strains in Manhiça district

From those 580 patients who started treatment during the study period in the district, 302 were microbiologically confirmed by culture, although 7 of those strains were not available for WGS due to poor quality of cultures. After exclusions (Figure 1), 275 strains were included in the analysis on population structure.

**Figure 1.**
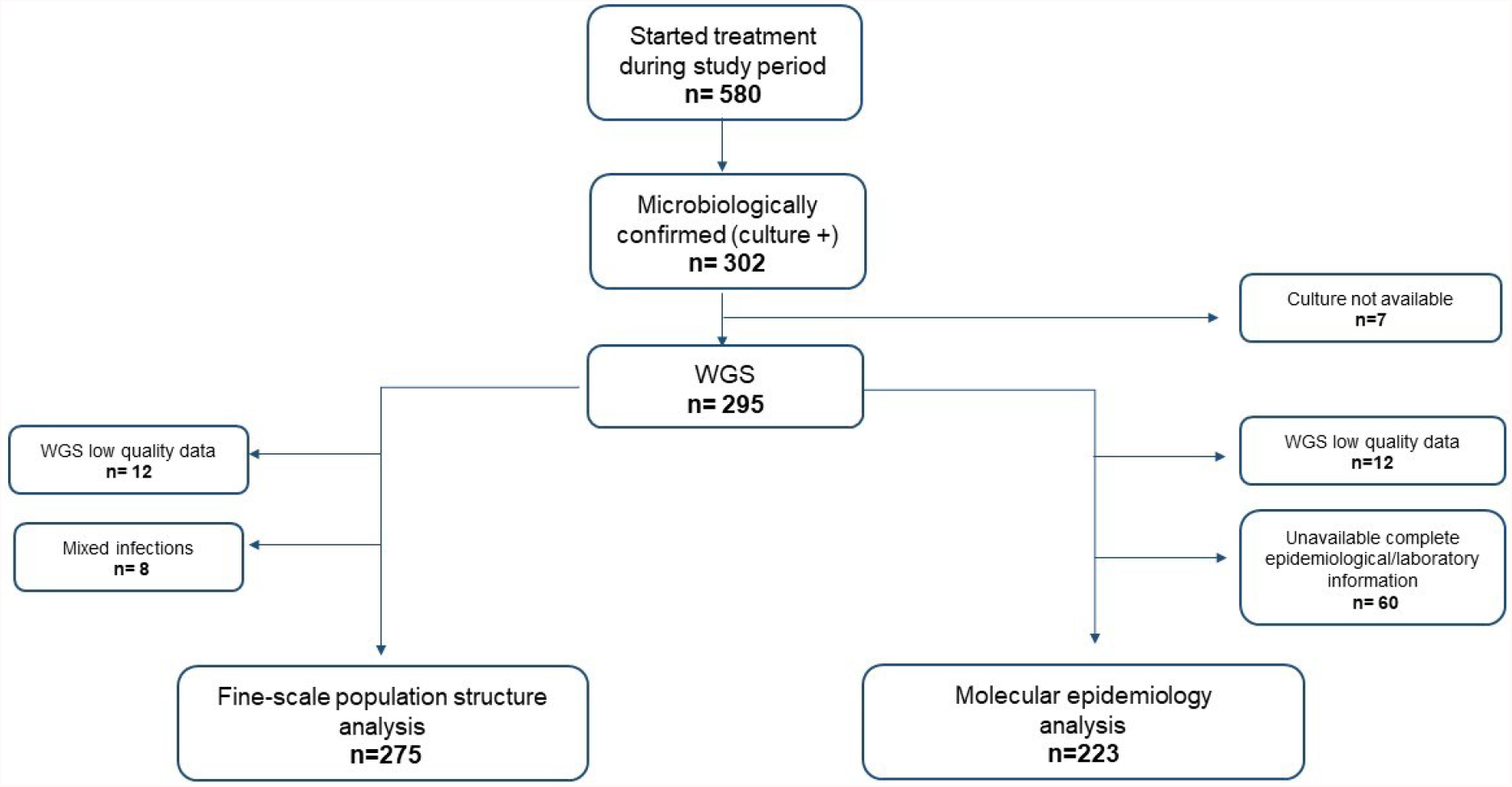
Study Flowchart. WGS: Whole genome sequencing

Overall, when reconstructing the phylogenetic tree (Figure 2), most cases (49.5%, 136/275) were classified as L4. The L4.3.4 (LAM) sub-lineage was the most common within L4 (23.3%, 64/275) although, at this level the most prevalent sublineage was L1.2 (25.4%, 70/275). The Beijing L2 were classified as L2.2.1, representing 12.7% (35/275) of the total population (Supplementary S1. Table 1). Additionally, eight samples were determined as mixed infections.

**Figure 2.**
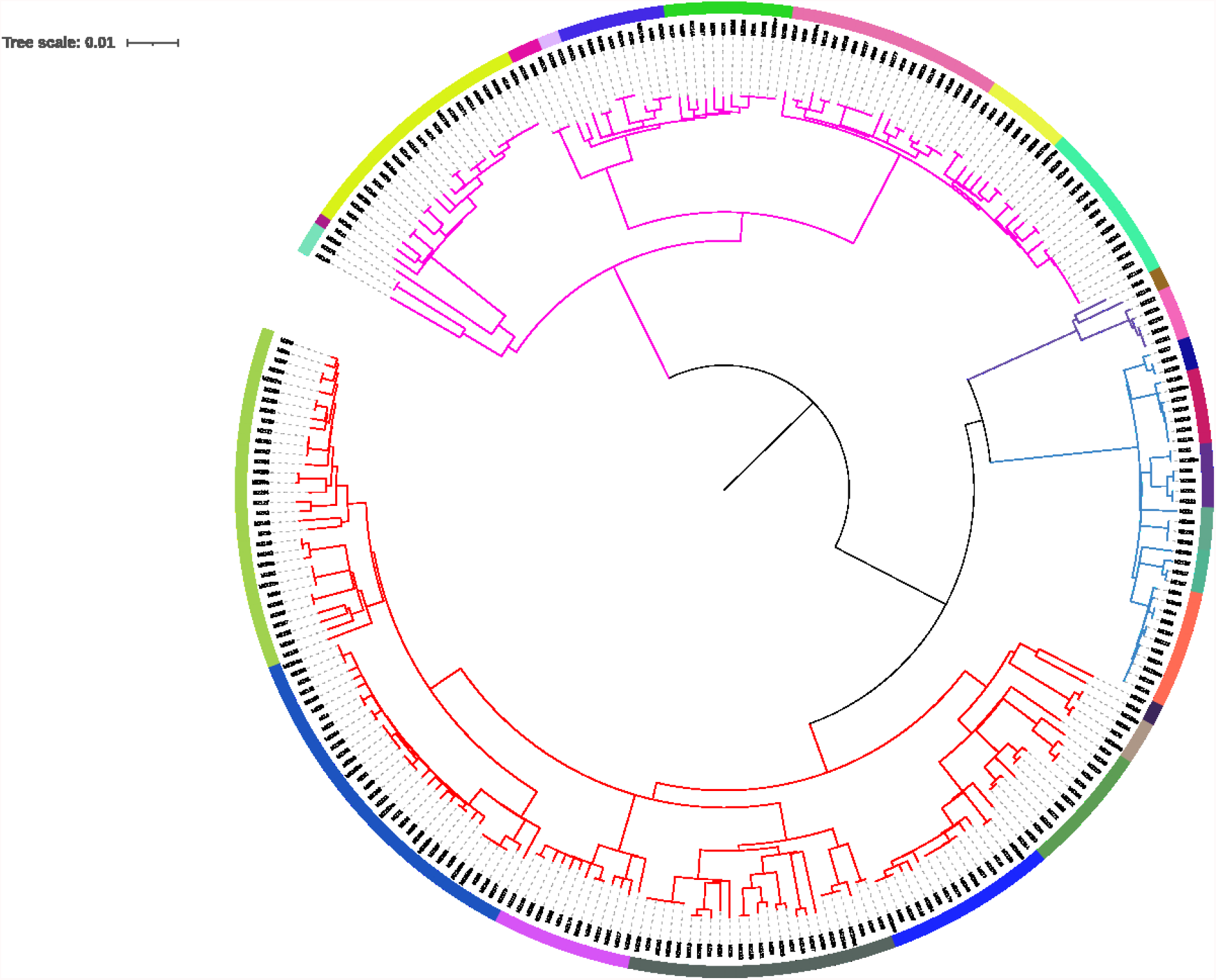
Phylogenetic tree of the 275 unique infections. Lineages are represented in branch colours (pink for L1, blue for L2, purple for L3 and red for L4). Twenty-five BAPS groups are denoted by different colours in the outer ring of the phylogeny.

### 2. SNPs-threshold approach to evaluate transmission events (Figure 4)

The clustering analysis revealed that 57.8% of strains (159/275), were in transmission, applying a cut-off of 5 SNPs. To evaluate the ‘transmission profile’ we compared genetic distances up to 10 SNPs with previously published data from Malawi (2009) and Valencia (2015) (See methods section). The distribution of pairwise distances revealed that, in all three regions, a large proportion of cases were in transmission, although patterns differed from one population to another, towering in Mozambique (64.7%) compared to Malawi (41.2%), and Valencia (29.8%).

While TB transmission seemed continuous for all three sites, the burden of very recent spread was significantly higher in Mozambique: 45.5% (125/275) of strains shared identical genotype (0 SNPs genetic distance), whereas only 9.2% (11/119) and 15.9% (41/246) were identified for Malawi and Valencia, respectively.

### 3. Fine-scale population structure of MTBC

By applying the BAPS algorithm, we recognized 25 groups of strains that shared high genomic similarity at level 2 of clustering hierarchy: 9 from L1, 6 L2, 2 L3, 8 L4 (1 strain did not assemble) (Figure 2). These groups were mapped in a global phylogeny (Figure 3) and, through the use of RASP, we inferred the probable geographic origin of the most recent common ancestor (MRCA) for each group (see methods).

**Figure 3.**
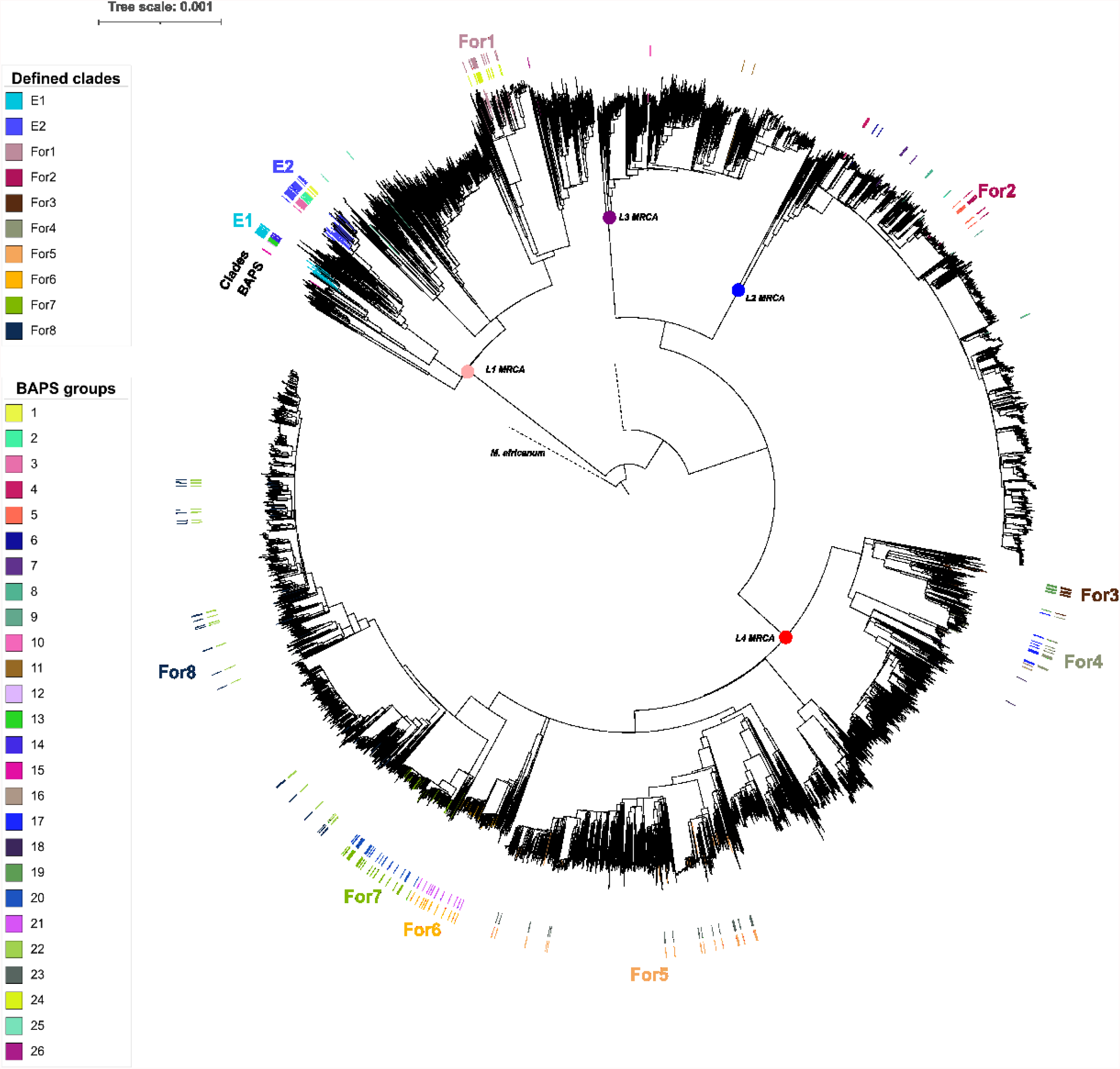
BAPS groups and inferred clades plotted in a global phylogeny. Footnotes: E: Endemic; For: Foreign (non-endemic); MRCA: Most recent common ancestror.

BAPS 1, 2, 3, 12, 13 and 14 were enriched in Mozambique isolates. BAPS 1 to 3, and 12 to 14 formed two largest groups sharing a MRCA with origin in the country (probability of 80%). Therefore, we merged those BAPS into two clades, and considered them ‘likely endemic’ or ‘local’ in our study setting (E1 and E2, 67/275, 24.4%) (Supplementary S2). Eight clades were defined as probably ‘non-endemic’ candidates (164 /275, 59.6%), and 11 did not accomplish defined criteria to be classified (see Methods). All strains categorized as endemic belonged to L1.2.

We then inferred the age of probable endemic and non-endemic clades (Supplementary S3). We found that, for those considered likely endemic, MRCAs located in Mozambique dated further back in time than in non-endemic ones. The mean of the median time when those local clades began to circulate in the country was 1894 [IQR 1887.7; 1900.6], whereas for non-endemic, it was estimated to be 2009 [IQR 1876.9; 2014.0] (Figure 4, Supplementary S4. Figure 4.1 & Figure 4.2).

**Figure 4.**
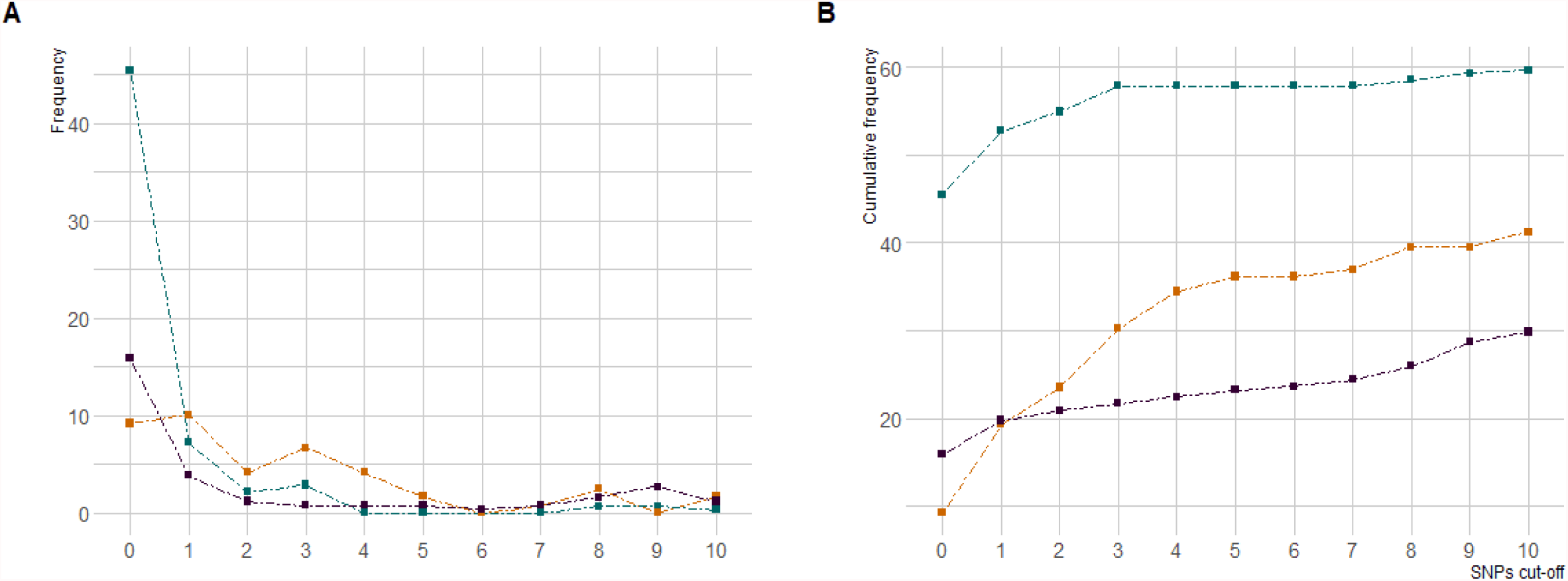
Proportion of strains linked by 0 to 10 SNPs. A) Frequency of samples found to each pairwise distance. B) Cumulative frequency (%) of isolates in transmission from 0 to 10 SNPs pairwise distance. Colours: the blue line represents data from Mozambique(2013-2014); orange from Malawi (2009) and purple from Valencia (2015).

**Figure 4.**
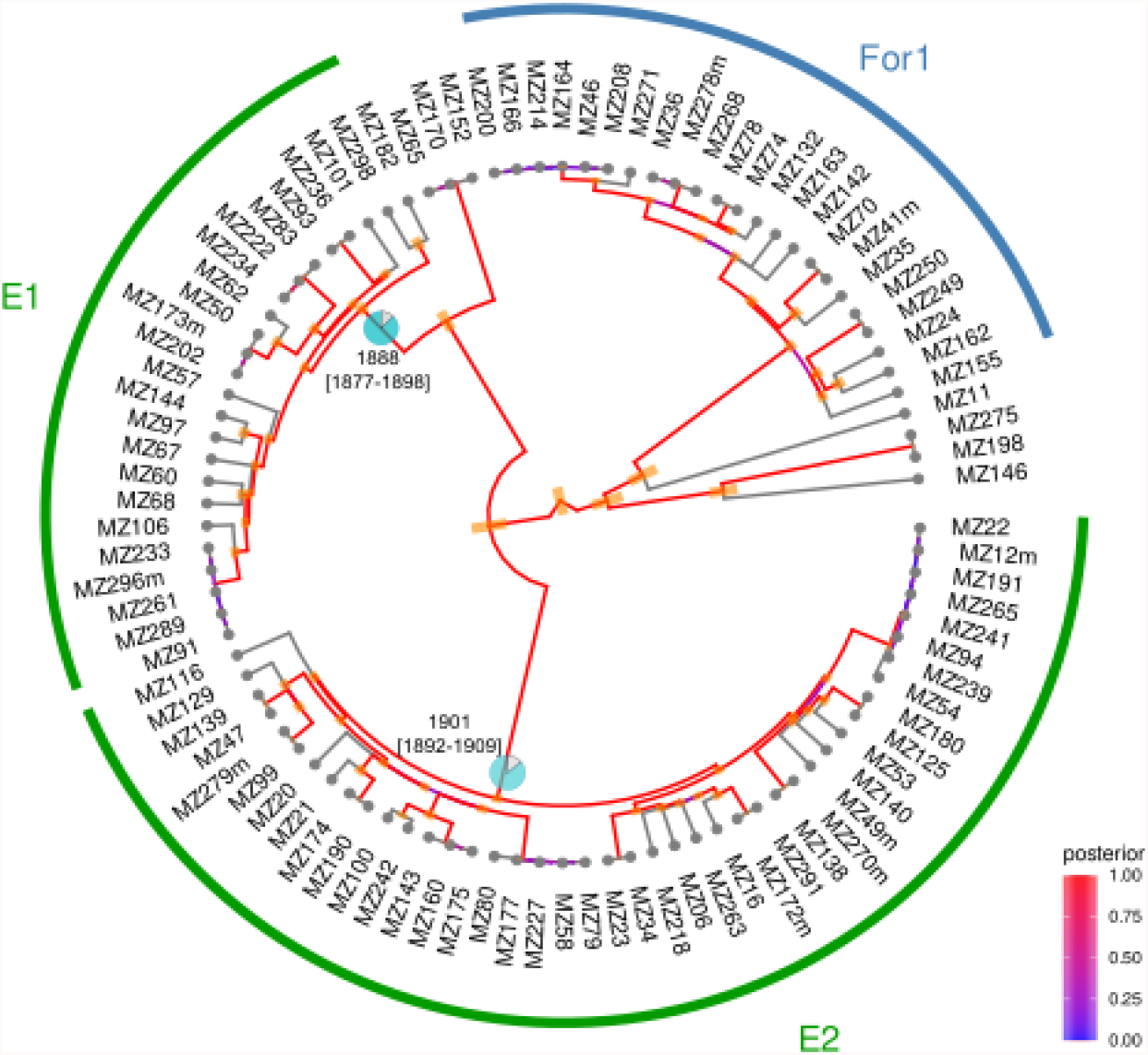
Endemic and Non-endemic clades from L1 with BEAST dating results for the MRCA with origin in Mozambique. Pie charts represent the probability that the geographical origin of the ancestor was MZ. The 95% Highest Posterior Density (HPD) is drawn as orange intervals. The posterior indicates the probability distribution over the parameter state space. The red colour (posterior close to one) indicates the maximum probability. Footnotes: E: Endemic; For: Foreign (non-endemic); 95% Highest Posterior Density

### 4. Impact of collected risk factor and endemicity on transmission

From those patients with WGS data, 223 strains had laboratory and epidemiological data available (Figure 1). These were used to investigate potential covariates associated with clustering, defined using a restrictive cut-off of pairwise distance at 5 SNPs to evaluate the burden of very recent transmission (see methods).

The median age of patients was 35 years [IQR 28.2;45.0], 35.4% (79/223) were female and the majority (72.2 %, 161/223) were people living with HIV (PLHIV). The median CD4+ count was 145.5 [49.9; 321.8] cells /mm3. The most common symptom was cough presented in 93.7% of cases (209/223). A sputum smear for acid-fast bacilli was positive in 65.0% (145/223) and 52.1% of participants had a pathological chest X-ray (CXR) (87/167 available). In this subset of data, 52.9% of isolates (118/223), were included in a cluster. Univariate exploration of other sociodemographic and microbiological variables is displayed in Table 1. The distribution of strains in recent transmission did not vary by sex, age, HIV status, symptoms, type of TB, different areas of the district or sublineage. Although none of the resistant strains were clustered, we were unable to explore this association further due to the limited number of isolates.

Based on the hypothesis that local genotypes might be linked to transmission, we explored the association between ‘endemicity’ and clustering. Overall, 24.2% (54/223) of strains were classified as ‘likely endemic’ (hereinafter endemic) and 28.0 % of them were in cluster (33/223). The results of the best fitted logistic regression model included are shown in Table 2. The odds of clustering did not differ by clades, whether endemic or not. Subsequent stratification by HIV status revealed that PLHIV who were infected with endemic strains had more than two times higher odds of being in recent transmission, regardless sex and age (2.11, 95%CI [1.04;4.45], p-value=0.04), and that this association disappeared among HIV-negative individuals.

Lastly, in light of those results, we investigated whether the degree of immunosuppression, measured through CD4 levels, might shape the distribution of endemic clades. We found that although the proportion of endemic strains increased as decreased the CD4 counts, this raise did not result meaningful (p-value=0.134) (Supplementary S5).

## Discussion

High resolution WGS data on MTBC population structure and transmission patterns is scarce in the country^13,43^, and similar to the situation for most high-burden countries. In order to provide a comprehensive insight on the molecular epidemiology of MTBC in Southern Mozambique, we have conducted the first population-based study using WGS data. The clustering analysis unmasked a high recent transmission profile. We also found two large likely-endemic clades that were associated with recent transmission among PLHIV. These clades were dated back to the end of the XIX century.

When pairwise distances were compared to one-year data from Malawi and Spain, profiles revealed that the highest proportion of strains involved in transmission was in Mozambique (64.7%)), and was also higher than that found in other high-burden settings (Liberia (39%, cut-off of 12 SNPs)^44^ and reported in Malawi for 15 years (31%).^40^ Although high rates of ongoing transmission were expected^3,45^, almost half of the strains were connected by 0 SNPs. Assuming a molecular clock of 0.3-0.5 SNPs/year, this would translate into very recent transmission events^46^ and fast rates of progression to active tuberculosis. Importantly, this proportion might be even underestimated as one-year population analyses are not enough to reveal the extent of recent transmission.

Interrupting ongoing spread is a critical public health intervention to bring down the current TB burden. Presented data reveals the suboptimal control of the epidemic in our setting and reinforce the need to explore the homegrown scenario. Most of participants were severely ill at the time of diagnosis, which might support previous works describing low awareness of disease^47,48^ and/or the existence of barriers in accessing medical care in our setting.^49^ Besides, programmatic active-case finding activities have been described as inconsistent in Mozambique, failing to reach hidden cases and breaking the transmission net.^50^ In our cohort of microbiologically-confirmed cases (highly transmitters), 20% of participants had contact with a TB case, although they sought health care because they were symptomatic (presumptive), proving the lack of active screening activities to identify patients in early stages.

The globally spread L4 was the most prevalent in our region, in agreement with previous works^10^, although at sublineage level, L1.2 was the most frequent. This finding is aligned with the global distribution of L1, concentrated around the Indian Ocean and that has been supposed to spread from South-Asia as a result of old migration pathways.^51^ To define clades specific to Mozambique,i.e. ‘probably endemic’, we applied a detailed, fine-scale population structure approach that allowed us to define two major endemic clades, both belonging to the L1.2 sub-lineage. In accordance to the hypothesis that they are endemic to our study setting, dating estimates confirmed they would have been circulating in Mozambique for longer times than non-endemic ones.

The long-term presence of those local strains in Mozambique suggests possible coevolution with the sympatric host population, as seen elsewhere.^52^ Thus, we tested the likelihood that endemic clades, as well as other collected covariates, could be related to a higher probability of recent spread using 5 SNPs as a threshold.^39^ Although through non-stratified models we did not find any remarkable result, posterior stratification by HIV status revealed that those classified as probably endemic presented higher odds of being responsible for recent transmission among HIV positive patients.

The extent to which TB/ HIV coinfection influences the structure of the MTBC population and the efficacy of transmission is currently under debate.^53,54^ On one hand, studies evaluating MTBC infectiousness among PLHIV are widely heterogeneous.^55,54^ Historically it has been argued that HIV decreases transmission, due to EPTB, lower cavitary lesions, or lower sputum bacillary load, among others.^53,56^ Nevertheless, recent works state that HIV-seropositive patients are as infectious as seronegative when they present cavitary disease or have a positive bacilloscopy.^54,57^ Furthermore, among PLHIV, most cases of active tuberculosis are the result of new infections.^58^ Therefore, for our setting, the large proportion of strains in recent spread may be due to the fact that half of PLHIV participants were severely immunosuppressed (highly susceptible to develop disease), microbiologically confirmed, and just 5% of them had extrapulmonary disease.

On the other hand, studies on coevolution have also demonstrated increased transmission between sympatric linage-host associations^11^, although, differently to our results, this was disrupted by HIV coinfections. Yet, the interdependence of both epidemics in high burden settings needs to embrace socioeconomical determinants that may shape the circulation and spread of MTBC and confound results. ^56^ Further studies on how HIV may influence sympatric associations in high burden areas are needed. Alternatively, our local population, regardless HIV status, may represent a reservoir for endemic strains with lower transmissibility, with some data suggesting that this is the case for the L1 specialist genotype.^59^ Limitations to understand this interplay includes the fact that the ancestry of individuals involved in this study is not known and that transmission estimates are based on a one-year sampling window. Although this may have limited our result, given the fast progression of the disease in PLHIV and the particularity of our dataset (45.5% of strains were 0 SNPs pairwise distance), one-year data may indeed be give us a hint of what is happening in our region.

Lastly, we cannot forget that host-related factors must be considered when interpreting genomic data. Information on how host and pathogen genotypes interplay is still scarce and few genotype-genotype interaction studies are available.^7^ Since the adaptive immune response is an essential mechanism for host recognition and control of MTBC^60^, we hypothesize that the interaction of HIV, MTBC and human populations is more complex in countries where the two epidemics collide with downstream impact on TB transmission. We highlight the need for population-based genome-to-genome association studies, including the MTBC, human and HIV genotype combined with sociodemographic data that could potentially confound results.

## Conclusion

Overall, our results unveil the population structure of MTBC in a high TB and HIV setting. We found an unexpected pattern of spread, with a substantial amount of strains of recent transmission, suggesting the uncontrolled TB spread and high rates of progression to active disease. We also identified endemic groups that were estimated to be circulating in the country for more than one century and that were responsible for recent transmission among PLHIV.

## Data Availability

Raw sequencing data will be available via online repository at the European Nucleotide Archive (ENA) under study accession number PRJEB27421 and SAMEA12029268.
De–identified dataset containing patient–level data produced in the present study are available upon reasonable request to the authors

## Contributors

All authors contributed as investigators to this study and to drafting and revising the final manuscript. All of them had full access to all the data in the study and had final responsibility for the decision to submit for publication.

## Acknowledgments

This project has received funding from the European Research Council (ERC) under the European Union’s Horizon 2020 research and innovation programs 101001038 (TB-RECONNECT), PID2019-104477RB-I00 from Ministerio de Economía y Competitividad (Spanish Government) (to I.C.).

We acknowledge support from the Spanish Ministry of Science, Innovation and Universities through the “Centro de Excelencia Severo Ochoa 2019-2023” Program (CEX2018-000806-S), and support from the Generalitat de Catalunya through the CERCA Program.

B.S receives a pre-doctoral fellowship from the Secretariat of Universities and Research, Ministry of Enterprise and Knowledge of the Government of Catalonia and co-funded by European Social Fund (AGAUR).

## Data availability

Raw sequencing data will be available via online repository at the European Nucleotide Archive (ENA) under study accession number PRJEB27421and SAMEA12029268. A limited de-identified dataset containing patient-level data will also be made available on publication.

## Supplementary material

- **Supplementary S1. Table 1. Distribution of lineages and sublineages overall and stratify by clustering and likely endemic clades**
- **Supplementary S2. Table 2. Summary of BAPs groups**
- **Supplementary S3. Table 3. Median time to the Mozambican Most Recent Common C (MRCA) for each clade and calculation of mean’s median times, by BAPS type (endemic (E) or foreign (For))**
- **Supplementary S4. Figure 4.1 BEAST tree for strains belonging to lineage 3; Figure 4.2. BEAST tree lineage 4**.
- **Supplementary S5. Figure S5. Distribution of endemic/non-endemic strains by HIV status, stratified by CD4 counts**.

## References

1 World Health Organization. Global Tuberculosis Report 2021. Geneva, Switzerland, 2021.

2 Gagneux S. Strain Variation in the Mycobacterium tuberculosis Complex: Its Role in Biology, Epidemiology and Control. 2017 DOI:10.1007/978-3-319-64371-7.

3 Yates TA, Khan PY, Knight GM, et al. The transmission of Mycobacterium tuberculosis in high burden settings. Lancet Infect Dis 2016; 16: 227–38.

4 Meehan CJ, Goig GA, Kohl TA, et al. Whole genome sequencing of Mycobacterium tuberculosis: current standards and open issues. Nat. Rev. Microbiol. 2019; 17: 533–45.

5 Comas I, Gagneux S. The Past and Future of Tuberculosis Research. PLoS Pathog 2009; 5: e1000600.

6 Brites D, Gagneux S. Old and new selective pressures on Mycobacterium tuberculosis. Infect Genet Evol 2012; 12: 678–85.

7 Gagneux S. Ecology and evolution of Mycobacterium tuberculosis. Nat Rev Microbiol 2018 164 2018; 16: 202–13.

8 Menardo F, Rutaihwa LK, Zwyer M, et al. Local adaptation in populations of Mycobacterium tuberculosis endemic to the Indian Ocean Rim. F1000Research 2021; 10: 60.

9 Freschi L, Vargas R, Husain A, et al. Population structure, biogeography and transmissibility of Mycobacterium tuberculosis. Nat Commun 2021 121 2021; 12: 1–11.

10 Stucki D, Brites D, Jeljeli L, et al. Mycobacterium tuberculosis lineage 4 comprises globally distributed and geographically restricted sublineages. Nat Genet 2016; 48: 1535–43.

11 Fenner L, Egger M, Bodmer T, et al. HIV Infection Disrupts the Sympatric Host-Pathogen Relationship in Human Tuberculosis. PLoS Genet 2013; 9. DOI:10.1371/journal.pgen.1003318.

12 Viegas SO, Machado A, Groenheit R, et al. Mycobacterium tuberculosis Beijing Genotype Is Associated with HIV Infection in Mozambique. PLoS One 2013; 8: 71999.

13 Namburete EI, Dippenaar A, Conceição EC, et al. Phylogenomic assessment of drug-resistant Mycobacterium tuberculosis strains from Beira, Mozambique. Tuberculosis 2020; 121: 1–6.

14 Sacoor C, Nhacolo A, Nhalungo D, et al. Profile: Manhica Health Research Centre (Manhica HDSS). Int J Epidemiol 2013; 42: 1309–18.

15 Nhacolo A, Jamisse E, Augusto O, et al. Cohort profile update: Manhiça health and demographic surveillance system (HDSS) of the Manhiça health research centre (CISM). Int J Epidemiol 2021; published online Jan 16. DOI:10.1093/ije/dyaa218.

16 Garcia-Basteiro AL, Hurtado JC, Castillo P, et al. Unmasking the hidden tuberculosis mortality burden in a large post mortem study in Maputo Central Hospital, Mozambique. Eur Respir J 2019; 54. DOI:10.1183/13993003.00312-2019.

17 García-Basteiro A, Ribeiro R, Brew J, et al. Tuberculosis on the rise in southern Mozambique (1997-2012). Eur Respir J 2017; 49: 1601683.

18 World Health Organization. Systematic screening for active tuberculosis. Principles and recommendations. Geneva, Switzerland, 2013 https://www.who.int/tb/tbscreening/en/ (accessed Aug 21, 2020).

19 Valencia S, Respeito D, Blanco S, et al. Tuberculosis drug resistance in Southern Mozambique: results of a population-level survey in the district of Manhiça. Int J Tuberc Lung Dis 2017; 21: 446–51.

20 Lumb R, Van Deun A, Bastian I, Fitz-Gerald M. Laboratory Diagnosis of Tuberculosis by Sputum Microscopy. The Handbook. Adelaide, South Australia, 2013.

21 Xpert MTB/RIF package insert. 2020.

22 Ssengooba W, Respeito D, Mambuque E, et al. Do xpert MTB/RIF cycle threshold values provide information about patient delays for tuberculosis diagnosis? PLoS One 2016; 11: 1–10.

23 Global Laboratory Initiative. Mycobacteriology Laboratory Manual. Geneva, Switzerland, 2014 https://www.who.int/tb/laboratory/mycobacteriology-laboratory-manual.pdf.

24 Illumina. Nextera XT DNA Library Prep Kit Reference Guide (15031942). 2019. www.illumina.com/company/legal.html. (accessed Dec 1, 2021).

25 TGU file repository – TUBERCULOSIS GENOMICS UNIT. http://tgu.ibv.csic.es/?page_id=1794 (accessed Dec 1, 2021).

26 Chen S, Zhou Y, Chen Y, Gu J. Fastp: An ultra-fast all-in-one FASTQ preprocessor. In: Bioinformatics. Oxford University Press, 2018: i884–90.

27 Wood DE, Salzberg SL. Kraken: Ultrafast metagenomic sequence classification using exact alignments. Genome Biol 2014; 15: 1–12.

28 Comas I. Genome of the inferred most recent common ancestor of the Mycobacterium tuberculosis complex. 2019; published online Oct 17. DOI:10.5281/ZENODO.3497110.

29 Li H, Durbin R. Fast and accurate long-read alignment with Burrows-Wheeler transform. Bioinformatics 2010; 26: 589–95.

30 Koboldt DC, Zhang Q, Larson DE, et al. VarScan 2: Somatic mutation and copy number alteration discovery in cancer by exome sequencing. Genome Res 2012; 22: 568–76.

31 Cingolani P, Platts A, Wang LL, et al. A program for annotating and predicting the effects of single nucleotide polymorphisms, SnpEff: SNPs in the genome of Drosophila melanogaster strain w1118; iso-2; iso-3. Fly (Austin) 2012; 6: 80–92.

32 Nguyen LT, Schmidt HA, Von Haeseler A, Minh BQ. IQ-TREE: a fast and effective stochastic algorithm for estimating maximum-likelihood phylogenies. Mol Biol Evol 2015; 32: 268–74.

33 Tonkin-Hill G, Lees JA, Bentley SD, Frost SDW, Corander J. RhierBAPs: An R implementation of the population clustering algorithm hierbaps [version 1; referees: 2 approved]. Wellcome Open Res 2018; 3: 93.

34 Yu Y, Harris AJ, Blair C, He X. RASP (Reconstruct Ancestral State in Phylogenies): A tool for historical biogeography. Mol Phylogenet Evol 2015; 87: 46–9.

35 Bouckaert R, Vaughan TG, Barido-Sottani J, et al. BEAST 2.5: An advanced software platform for Bayesian evolutionary analysis. PLOS Comput Biol 2019; 15: e1006650.

36 Menardo F, Duchêne S, Brites D, Gagneux S. The molecular clock of mycobacterium tuberculosis. PLoS Pathog 2019; 15: 1–24.

37 Rambaut A, Drummond AJ, Xie D, Baele G, Suchard MA. Posterior Summarization in Bayesian Phylogenetics Using Tracer 1.7. Syst Biol 2018; 67: 901–4.

38 Yu G, Smith DK, Zhu H, Guan Y, Lam TTY. ggtree: an r package for visualization and annotation of phylogenetic trees with their covariates and other associated data. Methods Ecol Evol 2017; 8: 28–36.

39 Walker TM, Lalor MK, Broda A, et al. Assessment of Mycobacterium tuberculosis transmission in Oxfordshire, UK, 2007-12, with whole pathogen genome sequences: An observational study. Lancet Respir Med 2014; 2: 285–92.

40 Guerra-Assuncao J, Crampin AC, Houben Rmgj, et al. Large-scale whole genome sequencing of M. tuberculosis provides insights into transmission in a high prevalence area. Elife 2015; 2015: 1–17.

41 Pennell MW, Eastman JM, Slater GJ, et al. geiger v2.0: an expanded suite of methods for fitting macroevolutionary models to phylogenetic trees. Bioinformatics 2014; 30: 2216–8.

42 Tonkin-Hill G, Lees JA, Bentley SD, Frost SDW, Corander J. Fast hierarchical Bayesian analysis of population structure. Nucleic Acids Res 2019; 47: 5539.

43 Viegas SO, MacHado A, Groenheit R, et al. Molecular diversity of Mycobacterium tuberculosis isolates from patients with pulmonary tuberculosis in Mozambique. BMC Microbiol 2010; 10: 195.

44 López MG, Dogba JB, Torres-Puente M, et al. Tuberculosis in Liberia: high multidrug-resistance burden, transmission and diversity modelled by multiple importation events. Microb Genomics 2020; 6. DOI:10.1099/mgen.0.000325.

45 Mathema B, Ph D, Ismail N, et al. Transmission of Extensively Drug-Resistant Tuberculosis in South Africa. 2017; : 243–53.

46 Meehan CJ, Moris P, Kohl TA, et al. The relationship between transmission time and clustering methods in Mycobacterium tuberculosis epidemiology. EBioMedicine 2018; 37: 410– 6.

47 Mindu C, López-Varela E, Alonso-Menendez Y, et al. Caretakers’ perspectives of paediatric TB and implications for care-seeking behaviours in Southern Mozambique. PLoS One 2017; 12: e0182213.

48 Noé A, Ribeiro RM, Anselmo R, et al. Knowledge, attitudes and practices regarding tuberculosis care among health workers in Southern Mozambique. BMC Pulm Med 2017; 17. DOI:10.1186/S12890-016-0344-8.

49 De Schacht C, Mutaquiha C, Faria F, et al. Barriers to access and adherence to tuberculosis services, as perceived by patients: A qualitative study in Mozambique. PLoS One 2019; 14. DOI:10.1371/JOURNAL.PONE.0219470.

50 José B, Manhiça I, Jones J, et al. Using community health workers for facility and community based TB case finding: An evaluation in central Mozambique. PLoS One 2020; 15. DOI:10.1371/JOURNAL.PONE.0236262.

51 Gagneux S. Host-pathogen coevolution in human tuberculosis. Philos. Trans. R. Soc. B Biol. Sci. 2012; 367: 850–9.

52 Gagneux S, DeRiemer K, Van T, et al. Variable host-pathogen compatibility in Mycobacterium tuberculosis. Proc Natl Acad Sci U S A 2006; 103: 2869–73.

53 Africa S Peters SJ, Kana BD, Peters JS, et al. Tuberculosis transmission in HIV-endemic settings 1 Advances in the understanding of Mycobacterium tuberculosis transmission in HIV-endemic settings. Lancet Infect Dis 2019; 19: e65–76.

54 Martinez L, Woldu H, Chen C, et al. Transmission Dynamics in Tuberculosis Patients With Human Immunodeficiency Virus: A Systematic Review and Meta-analysis of 32 Observational Studies. Clin Infect Dis An Off Publ Infect Dis Soc Am 2021; 73: e3446.

55 Kendall EA, Kendall EA. When Infections Don’t Reflect Infectiousness: Interpreting Contact Investigation Data With Care. Clin Infect Dis 2021; 73: e3456–8.

56 Kwan C, Ernst JD. HIV and Tuberculosis: a Deadly Human Syndemic. Clin Microbiol Rev 2011; 24: 351.

57 Martinez L, Sekandi JN, Castellanos ME, Zalwango S, Whalen CC. Infectiousness of HIV-seropositive patients with tuberculosis in a high-burden African Setting. Am J Respir Crit Care Med 2016; 194: 1152–63.

58 Cudahy PGT, Andrews JR, Bilinski A, et al. Spatially targeted screening to reduce tuberculosis transmission in high-incidence settings. Lancet Infect Dis 2019; 19: e89–95.

59 Freschi L, Vargas R, Husain A, et al. Population structure, biogeography and transmissibility of Mycobacterium tuberculosis. Nat Commun 2021 121 2021; 12: 1–11.

60 Comas I, Chakravartti J, Small PM, et al. Human T cell epitopes of Mycobacterium tuberculosis are evolutionarily hyperconserved. Nat Genet 2010; 42: 498–503.

