## Supplementary material for "Unravelling the population structure and transmission patterns of *Mycobacterium tuberculosis* in Mozambique, a high TB/HIV burden country"

**Supplementary S1. Table 1. Distribution of lineages and sublineages overall and stratify by clustering and likely endemic clades**

| **Lineage** | **n= 275** | **(%)** | | |
|---|---|---|---|---|
| 1 | 97 | 35.3 | | |
| 2 | 35 | 12.7 | | |
| 3 | 7 | 2.5 | | |
| 4 | 136 | 49.5 | | |
| **Sublineages** | | | | |
| L1.1 | 27 | 9.8 | | |
| L1.2 | 70 | 25.4 | | |
| L2.2.1 | 35 | 12.7 | | |
| L3 | 7 | 2.6 | | |
| L4.1 | 34 | 12.4 | | |
| L4.3.4 | 64 | 23.3 | | |
| L4.4 | 13 | 4.7 | | |
| L4.10 | 25 | 9.1 | | |
| **Sublineages[1]** | **Unclustered (116)** | **(%)** | **Clustered (159)** | **(%)** |
| L1.1 | 10 | 8.6 | 17 | 10.7 |
| L1.2 | 25 | 21.5 | 45 | 28.3 |
| L2.2 | 9 | 7.8 | 26 | 16.3 |
| L3 | 4 | 3.4 | 3 | 1.9 |
| L4.1 | 16 | 13.8 | 18 | 11.3 |
| L4.3 | 32 | 27.6 | 32 | 20.1 |
| L4.4 | 9 | 7.8 | 4 | 2.5 |
| L4.10 | 11 | 9.5 | 14 | 8.8 |
| **Sublineages[2]** | **Non-endemic (208)** | **(%)** | **Endemic (67)** | **(%)** |
| L1.1 | 27 | 13.0 | 0 | 0 |
| L1.2 | 3 | 1.4 | 67 | 100 |
| L2.2 | 35 | 16.8 | 0 | 0 |
| L3 | 7 | 3.4 | 0 | 0 |
| L4.1 | 34 | 16.3 | 0 | 0 |
| L4.3 | 64 | 30.8 | 0 | 0 |
| L4.4 | 13 | 6.2 | 0 | 0.0 |
| L4.10 | 25 | 12.0 | 0 | 0.0 |

[1] p-value=0.09 ; [2] p-value<0.001 Fischer-exact test

**Supplementary S2. Table 2. Summary of BAPs groups**

| BAPS group | Type of clade | lineage | clade name | Number of strains |
|---|---|---|---|---|
| 1 | endemic | L1 | E2 | 8 |
| 2 | endemic | L1 | E2 | 15 |
| 3 | endemic | L1 | E2 | 20 |
| 4 | unknown | L2 | Unk4 | 7 |
| 5 | non-endemic | L2 | For2 | 11 |
| 6 | unknown | L2 | Unk5 | 3 |
| 7 | unknown | L2 | Unk6 | 6 |
| 8 | unknown | L2 | Unk7 | 4 |
| 9 | unknown | L2 | Unk8 | 4 |
| 10 | unknown | L3 | Unk10 | 5 |
| 11 | unknown | L3 | Unk11 | 2 |
| 12 | endemic | L1 | E1 | 2 |
| 13 | endemic | L1 | E1 | 12 |
| 14 | endemic | L1 | E1 | 10 |
| 15 | unknown | L1 | Unk3 | 3 |
| 16 | unknown | L4 | Unk10 | 4 |
| 17 | foreign | L4 | For4 | 16 |
| 18 | unknown | L4 | Unk9 | 2 |
| 19 | non-endemic | L4 | For3 | 12 |
| 20 | non-endemic | L4 | For7 | 32 |
| 21 | non-endemic | L4 | For6 | 13 |
| 22 | non-endemic | L4 | For8 | 33 |
| 23 | non-endemic | L4 | For5 | 25 |
| 24 | non-endemic | L1 | For1 | 23 |
| 25 | unknown | L1 | Unk1 | 3 |
| 26 | unknown | L1 | Not grouped | 1 |

**Footnotes:** E: Endemic; For: Foreign (non-endemic); Unk: Unknown, not possible to be classified

**Supplementary S3. Table 3. Median time to the Mozambican Most Recent Common C (MRCA) for each clade and calculation of mean´s median times, by BAPS type (endemic(E) or foreign (For))**

| | | | Estimated time of the Mozambican MRCA | |
|---|---|---|---|---|
| **BAPS type** | **Lineage** | **Clade** | **Median time (years)** | **95% HPD[1] (years)** |
| endemic | L1 | E1 | 1887.73 | [1877.4 – 1898] |
| endemic | L1 | E2 | 1900.61 | [1891.9 – 1909.1] |
| **Mean Endemic** | | | **1894.17** | **Range [1887.7 – 1900.6]** |
| foreign | L1 | For1-1 | 1932.45 | [1923.6 – 1941.1] |
| foreign | L1 | For1-2 | 2014 | |
| foreign | L1 | For1-3 | 2014 | |
| foreign | L1 | For1-4 | 2014 | |
| foreign | L1 | For1-5 | 2013.57 | [2012.2 – 2014] |
| foreign | L1 | For1-6 | 2014 | |
| foreign | L1 | For1-7 | 2013.62 | [2012.5 – 2014] |
| foreign | L2 | For2-1 | 2013.22 | [2010.5 – 2014] |
| foreign | L2 | For2-2 | 1982.07 | [1966.5 – 1995.4] |
| foreign | L2 | For2-3 | 2014 | |
| foreign | L4 | For3-1 | 2012.76 | [2008 – 2014] |
| foreign | L4 | For3-2 | 2014.00 | |
| foreign | L4 | For3-3 | 2014.00 | |
| foreign | L4 | For3-4 | 2014.00 | |
| foreign | L4 | For3-5 | 2014.00 | |
| foreign | L4 | For3-6 | 2014.00 | |
| foreign | L4 | For3-7 | 2012.89 | [2009.2 – 2014] |
| foreign | L4 | For3-8 | 2012.82 | [2009.2 – 2014] |
| foreign | L4 | For4-1 | 2008.84 | [2002.7 – 2013.3] |
| foreign | L4 | For4-2 | 2005.50 | [1998.2 – 2011.3] |
| foreign | L4 | For4-3 | 2012.79 | [2008.7 – 2014] |
| foreign | L4 | For4-4 | 2014 | |
| foreign | L4 | For4-5 | 2014 | |
| foreign | L4 | For4-6 | 2014 | |
| foreign | L4 | For4-7 | 2014 | |
| foreign | L4 | For4-8 | 2014 | |
| foreign | L4 | For5-1 | 1917.98 | [1885.4 – 1947.3] |
| foreign | L4 | For5-2 | 2011.43 | [2006.2 – 2014] |
| foreign | L4 | For5-3 | 2008.65 | [2002.5 – 2013.1] |
| foreign | L4 | For5-4 | 2012.57 | [2007.8 – 2014] |
| foreign | L4 | For5-5 | 2010.99 | [2005.4 – 2014] |
| foreign | L4 | For5-6 | 2011.14 | [2005.4 – 2014] |
| foreign | L4 | For5-7 | 2014 | |
| foreign | L4 | For5-8 | 2014 | |
| foreign | L4 | For5-9 | 2014 | |
| foreign | L4 | For5-10 | 2014 | |

| | | | | |
|---|---|---|---|---|
| foreign | L4 | For5-11 | 2014 | |
| foreign | L4 | For5-12 | 2014 | |
| foreign | L4 | For5-13 | 2014 | |
| foreign | L4 | For5-14 | 2014 | |
| foreign | L4 | For5-15 | 2014 | |
| foreign | L4 | For6-1 | 2013.07 | [2009.1 – 2014] |
| foreign | L4 | For6-3 | 2014 | |
| foreign | L4 | For6-4 | 2014 | |
| foreign | L4 | For6-5 | 2014 | |
| foreign | L4 | For6-6 | 2014 | |
| foreign | L4 | For6-7 | 2014 | |
| foreign | L4 | For6-8 | 2014 | |
| foreign | L4 | For6-9 | 2014 | |
| foreign | L4 | For6-10 | 2014 | |
| foreign | L4 | For6-11 | 2014 | |
| foreign | L4 | For7-1 | 1998.36 | [1987.1 – 2007.2] |
| foreign | L4 | For7-2 | 2008.97 | [2002.8 – 2012.8] |
| foreign | L4 | For7-3 | 2008.33 | [2001.7 – 2012.7] |
| foreign | L4 | For7-4 | 2007.93 | [2000.7 – 2012.6] |
| foreign | L4 | For7-5 | 1876.90 | [1852.9 – 1899.6] |
| foreign | L4 | For7-6 | 2011.74 | [2007 – 2013.8] |
| foreign | L4 | For7-7 | 2008.95 | [2002.6 – 2013.1] |
| foreign | L4 | For7-8 | 2012.72 | [2007.9 – 2014] |
| foreign | L4 | For7-9 | 2014 | |
| foreign | L4 | For7-10 | 2014 | |
| foreign | L4 | For7-11 | 2014 | |
| foreign | L4 | For7-12 | 2014 | |
| foreign | L4 | For7-13 | 2014 | |
| foreign | L4 | For7-14 | 2014 | |
| foreign | L4 | For7-15 | 2014 | |
| foreign | L4 | For7-16 | 2014 | |
| foreign | L4 | For7-17 | 2014 | |
| foreign | L4 | For7-18 | 2014 | |
| foreign | L4 | For8-1 | 1988.60 | [1973.5 – 2000.9] |
| foreign | L4 | For8-2 | 2004.50 | [1995.7 – 2010.5] |
| foreign | L4 | For8-3 | 2011.88 | [2007.2 – 2014] |
| foreign | L4 | For8-4 | 2012.88 | [2008.8 – 2014] |
| foreign | L4 | For8-5 | 2013 | [2009.4 – 2014] |
| foreign | L4 | For8-6 | 2011.33 | [2005.9 – 2014] |
| foreign | L4 | For8-7 | 2012.59 | [2014 – 2012.6] |
| foreign | L4 | For8-8 | 2014 | |
| foreign | L4 | For8-9 | 2014 | |
| foreign | L4 | For8-10 | 2014 | |
| foreign | L4 | For8-11 | 2014 | |
| foreign | L4 | For8-12 | 2014 | |
| foreign | L4 | For8-13 | 2014 | |

| | | | | |
|---|---|---|---|---|
| foreign | L4 | For8-14 | 2014 | |
| foreign | L4 | For8-15 | 2014 | |
| foreign | L4 | For8-16 | 2014 | |
| foreign | L4 | For8-17 | 2014 | |
| foreign | L4 | For8-18 | 2014 | |
| foreign | L4 | For8-19 | 2014 | |
| foreign | L4 | For8-20 | 2014 | |
| foreign | L4 | For8-21 | 2014 | |
| foreign | L4 | For8-22 | 2014 | |
| foreign | L4 | For8-23 | 2014 | |
| **mean Foreign** | | | **2008.92** | **Range [1876.9 – 2014]** |

**Footnote: [1] 95% HPD: Highest Posterior Density**

**Supplementary S4.**

**Figure 4.1 BEAST tree for strains belonging to lineage 3**

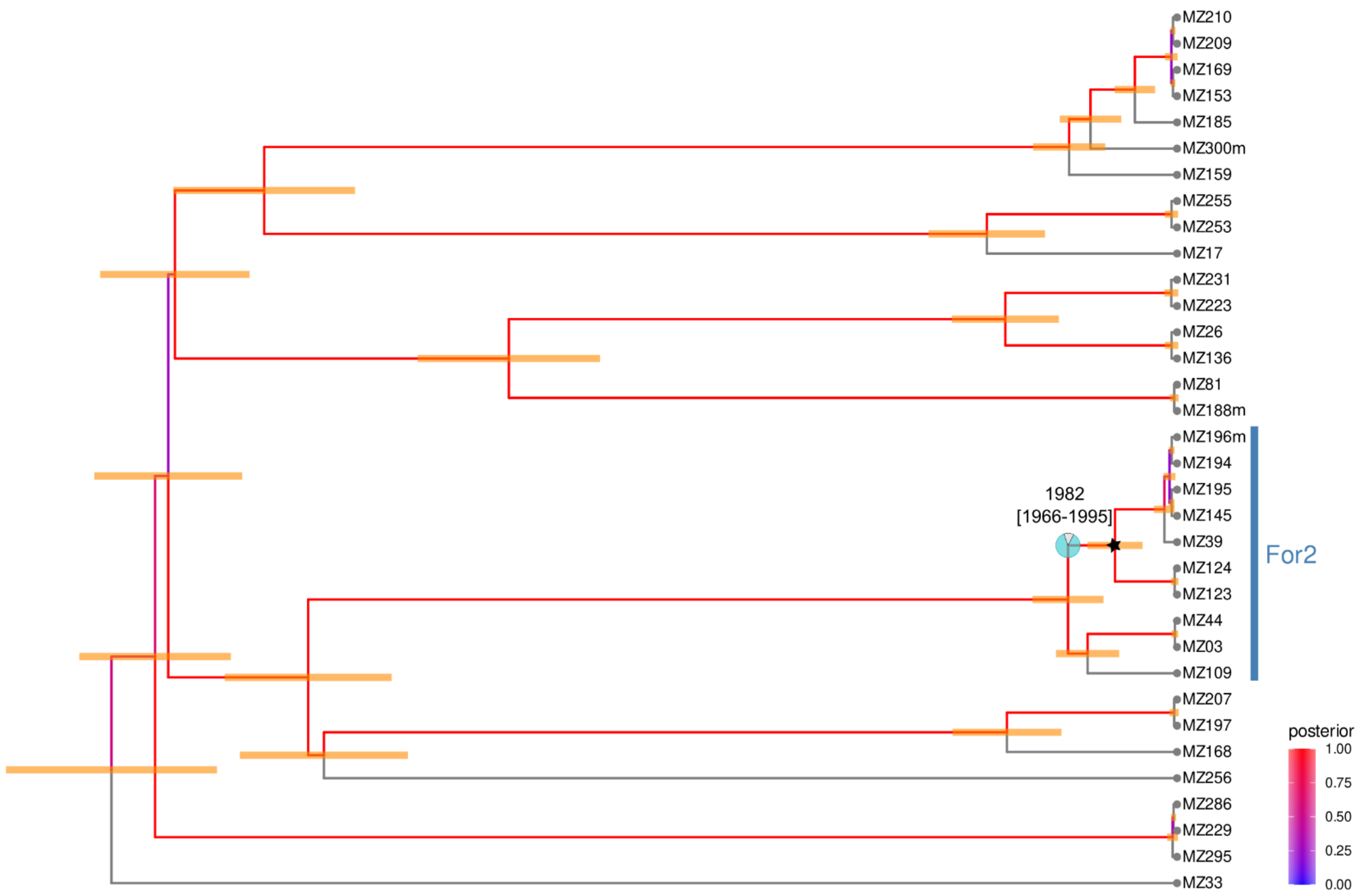


**L3 BEAST tree. Dating results for the MRCA with origin in Mozambique are displayed.** Pie charts represents the probability that the geographical origin of the ancestor was MZ. The 95% HPD is drawn as orange intervals. The posterior indicates the probability distribution over the parameter state space. The red colour (posterior close to one) indicates the maximum probability. For: Foreign

**Figure 4.2. BEAST tree lineage 4**

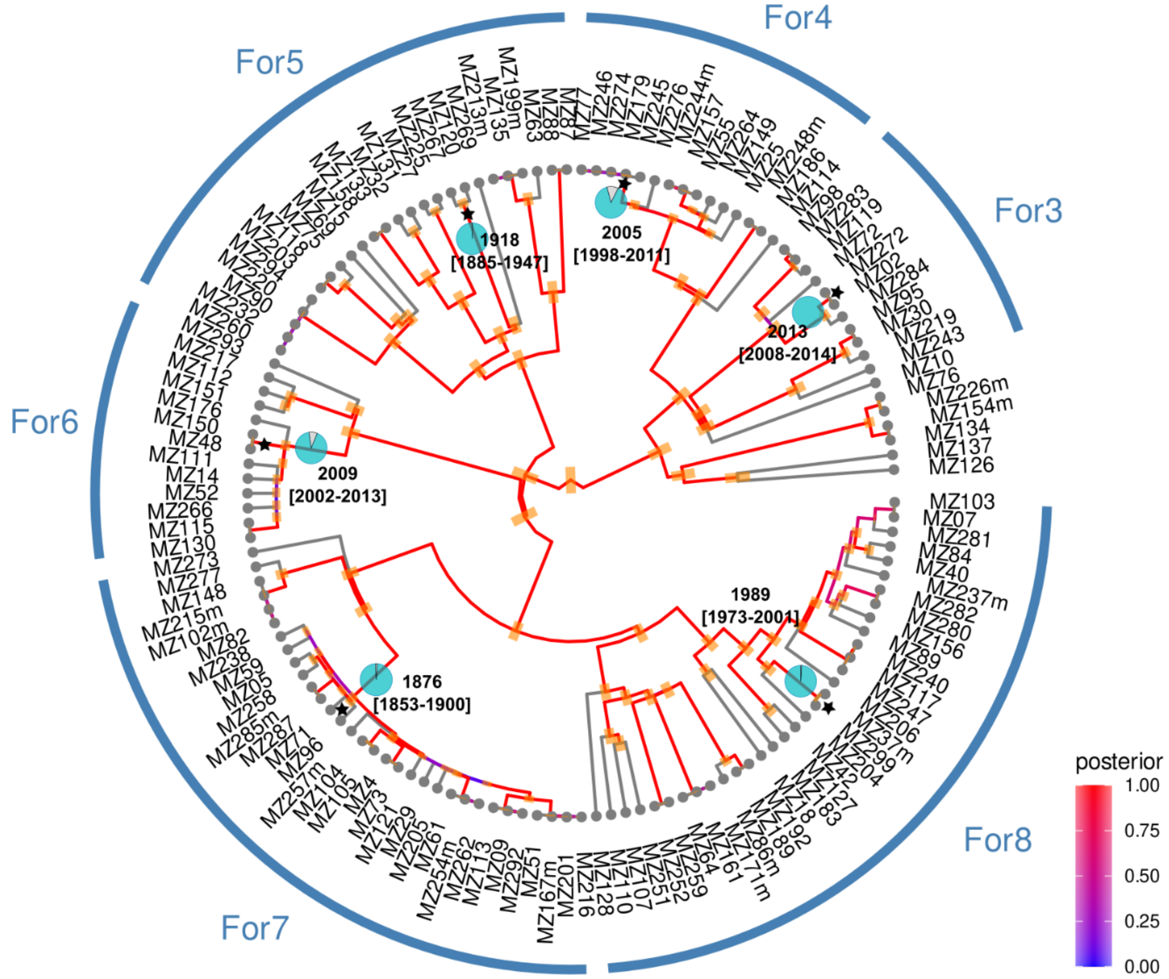


**L4 BEAST tree. Dating results for the MRCA with origin in Mozambique are displayed.** Pie charts represents the probability that the geographical origin of the ancestor was MZ. The 95% HPD is drawn as orange intervals. The posterior indicates the probability distribution over the parameter state space. The red colour (posterior close to one) indicates the maximum probability.

Footnotes: E: Endemic; For: Foreign

**Supplementary S5.** Graphical representation of relation between immunity status of participants and endemicity and data on the proportion of endemic and non-endemic clades stratified by CD4 counts  PLHIV: People living with HIV

| | **Endemic (%)** [2] | **Non-endemic (%)** | **p-value**[3] |
|---|---|---|---|
| **(n=188)** [1] | | | 0·130 |
| **>200 cc/mm3** | 14 (25·9) | 40 (74·1) | |
| **<200 cc/mm3** | 25 (31·6) | 54 (68·4) | |
| **Negative** | 9 (16·4) | 46 (83·6) | |

**Footnote**: [1]excluding coinfections (8), HIV-status missing (2) and CD4 missing (25); [2]row percentages; [3] Fisher´s exact tes

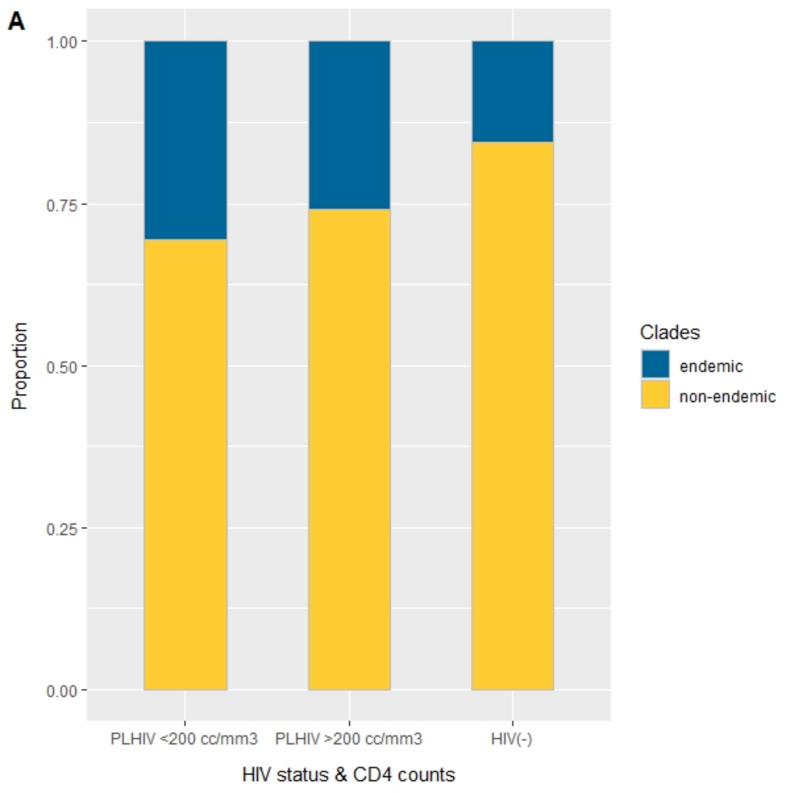
